# Reproductive number of COVID-19: A systematic review and meta-analysis based on global level evidence

**DOI:** 10.1101/2020.05.23.20111021

**Authors:** Md. Arif Billah, Md. Mamun Miah, Md. Nuruzzaman Khan

## Abstract

**Background:** The coronavirus (COVID-19) is now a global concern because of its higher transmission capacity and associated adverse consequences including death. The reproductive number of COVID-19 provides an estimate on possible extent of the transmission. This study aims to provide the average reproductive number of COVID-19 based on available global level evidence.

**Methods:** We searched three databases (PubMed, Web of Science, and Science Direct) to find studies reported the reproductive number of COVID-19. The searches were conducted using a pre-specified search strategy that includes keywords of COVID-19 and its reproductive number related terms, which were combined using the Boolean operators. We used meta-analysis to provide average reproduction number of COVID-19.

**Results:** Total of 30 studies included in this review whereas 24 of them were included in the meta-analysis. The average estimated reproductive number was 2.70 (95% CI, 2.21-3.30). We found evidence of very high heterogeneity (99.5%) of the reproductive number reported in the included studies. The highest reproductive number was reported for Diamond Princes Cruise Ship, Japan (14.8). In the country-level, higher reproductive number was reported for France (R, 6.32, 95% CI, 5.72-6.98) following Germany (R, 6.07, 95% CI, 5.51-6.69) and Spain (R, 5.08, 95% CI, 4.50-5.73). We also found estimation models, methods, and the number of cases considered to estimate reproductive number were played a role in arising the heterogeneity of the estimated reproductive number.

**Conclusion:** The estimated reproductive number indicates an exponential increase of COVID-19 infection in coming days. Comprehensive policies and programs are important to reduce new infections as well as the associated adverse consequences including death.

## Background

Coronavirus (COVID-19) is now a global concern that speared out to 213 countries or territories as of May 30, 2020. More than 6 million population have been infected so far worldwide, of which more than 367,304 are died [1]. Consequently, the World Health Organization has declared it as pandemic and suggested countries to take aggressive measures to reduce new infections [2]. Given no treatments or vaccines available for this virus, countries are now also taking numerous non-medical measures to reduce further infections, which include restricting people’s movements, banned international and local travels, quarantine, and isolation [3]. However, the new infections are rising exponentially, in all ages and sexes, irrespective of countries [4,5]. Reducing new infections, therefore, needs further comprehensive preventive measures.

The virus was first discovered by Tyrrell and Bynoe in 1965 in the human respiratory tract of an adult infected with the common cold [6]. Since then the virus caused three major large-scale outbreaks, namely, Severe Acute Respiratory Syndrome (SARS) in 2003 in mainland China [7], Middle East Respiratory Syndrome (MERS) in 2012 in Saudi-Arabia [8], and MERS in 2015 in South Korea [9]. These outbreaks showed some similar characteristics which are common with current COVID-19 outbreak, such as fever, cough, and the breathing difficulties [10].

This round of coronavirus appeared from a single center in a seafood market of Wuhan, China [11]. Two families consisting of five people or healthcare workers infected initially from the seafood market and visited a nearest healthcare center, from where the virus was spread rapidly to other people (nosocomial) through human-to-human transmission [12-16]. The virus then spread out worldwide through international travelers from China (Figure 1) [17].

**Figure 1:**
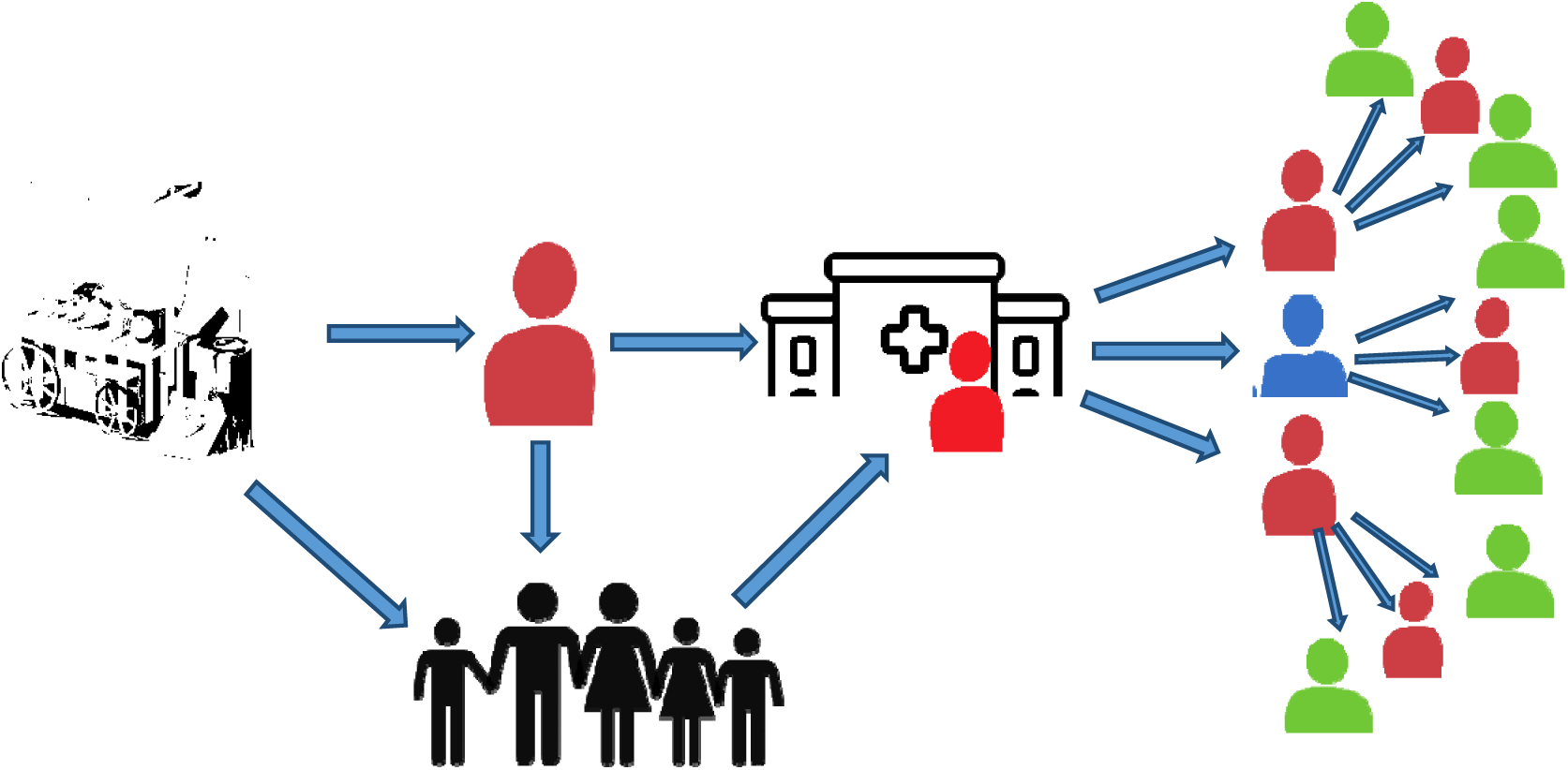
The probable ways of the COVID-19 transmission into the human and turned into a person-to-person transmission process (The seafood shop was the center from where the virus transmitted into customers or sellers or both. The symptoms of the infection were normal cold, headache, and pneumonia. The infected persons then transmitted the virus to non-infected in two different routes: (a). Infected persons went to the nearby hospital for treatment. Clinical manifestations were not examined properly and the infected persons presented risk to transmission among non-infected hospital attendances (b). Infection also transmitted into the infected person family members. The virus was then spread out worldwide, at the end of February 2020, through the infected international travelers from China).

Knowing the accurate reproductive number of COVID-19, which means the capability of transmission per primary infected person to the secondarily infected persons, is significant for various reasons: to assess epidemic transmissibility and to predict the future trend of spreading [18]. These are important to reduce new infections and to design control measures such as social distancing [19] and to know the duration of keeping control measures [5]. Moreover, it also helps to develop an effective epidemiological mathematical models considering possible transmission ways, such as, droplets and direct contacts with COVID-19 infected people, which are important to know the risk population and the appropriate epidemiologic parameters [20,21].

Various researchers worldwide estimated reproductive number of COVID-19. However, these were not consistent and measurement procedures and methods were different across the studies [20,22]. The reproductive number estimated was also found different across the countries, stages of infection, and the preventive measures applied [23]. Another important source of variation of estimated reproductive number was type of reproductive numbers considered [20]. Of the three reproductive numbers estimated, namely basic reproductive number (*R*_0_), net reproductive number (*R_e_*), and time dependent reproductive number (*R_t_*), are applicable for different purposes. For instance, the basic reproductive number is used when an infected person can mix randomly to non-infected persons (i.e no control intervention was applied), whereas, the net and time-dependent reproductive number are used when control interventions were applied. Consequently, these three reproductive numbers are also followed different distributions of infection period. However, the value of each reproductive number ranges from zero to any positive number, where *R* < 1 indicates new infection will decrease, *R* < 0 indicates the stability of new infection, and *R* < 1 indicates new infection will increase [24,25].

Considering the higher variability of the reproductive number estimated and its underlying importance, in this study, an attempt has been made to summarize available reproductive number of COVID-19 to give an average estimate. If applicable, sources of variations of the estimated reproductive number will also be addressed. Findings will help policymakers to know about the possible increase of COVID-19’s patients and take policies and programs accordingly.

## Methods

Literature searches were conducted in three databases on April 10, 2020: PubMed, Web of Science, and Science Direct. The pre-specified search strategies were used to search databases (Supplementary Tables 1-3). We developed search strategies consisting of virus-specific (corona virus, coronavirus, SARS-CoV-2, COVID-19, nCoV-2019) and reproductive number related (reproduction number, transmissibility) keywords that were combined using the Boolean operators (AND, OR). Additional searches were conducted in the reference list of the selected articles, and the relevant journal’s websites.

### Inclusion and exclusion criteria

Studies meet the following inclusion criteria were included: wrote in the English language, related to COVID-19, and presented the reproductive number of COVID-19. We did not apply any time restriction, i.e. all studies from the onset of COVID-19 to the date of conducting formal search were included. Studies that did not meet these criteria were excluded.

### Data extraction and analysis

Two authors (MAB, MMM) extracted information by using a pre-designed, trailed, and modified data extraction sheet. The extracted information includes: year of publication, study’s location, model used to estimate the reproductive number, time period for when the reproductive number was estimated, number of cases considered to estimate the reproductive number, assumption(s) that was/were set to a calculate the reproductive number, intervention strategy, and the estimated reproductive number with its 95% confidence interval (CI). The corresponding author (MNK) solved any disagreement on information extraction.

The information recorded were mostly dichotomous in nature where the numerical reproductive number was reported in all selected studies. We, therefore, used both narrative synthesis and meta-analysis to summaries findings from retrieved studies. Narrative synthesis used initially to describe assumptions applied in estimating the reproductive number, number of cases used to estimate reproductive number, time/period for when the reproductive number estimated, and the models and methods used to estimate the reproductive number. Meta-analysis then used to give an average estimate of the reproductive number. We used both fixed effect and random effect model to summarize the reproductive number selected based on heterogeneity assessment (*I^2^*). We used fixed effect model if the heterogeneity was low (*I*^2^ < 50%) and the random effect model if the heterogeneity was moderate (*I*^2^ < 50%) or high (*I*^2^ < 75%). For the studies where more than one reproductive number reported based on different assumptions, we calculated the average reproductive number. This calculated average reproductive number was then used to give summary estimate of the reproductive number. Heterogeneity of the average estimated reproductive number was assessed through sub-groups analysis across the selected studies’ characteristics. We also assessed the publication bias through visual inspection of the funnel plot and Egger’s regression asymmetry test. Trim-and-Fill procedure was used when evidence of publication bias was found. The National Institutes of Health (NIH) study quality assessment tool was used to assess study quality. The Stata software version 15.1 (Stata Corp, College Station, Texas, USA) was used to perform all analyses.

## Results

### Literature search results

Total of 134 studies included, 130 of them were extracted from three databases searched (Figure 2, Supplementary Tables 1-3). Of these, 102 studies were excluded through title and abstract screening leaving 32 studies for full-text review. A total of 30 of them were finally included in this study and 24 of them were included in meta-analysis. All study were high in quality (Supplementary Table 4).

**Figure 2.**
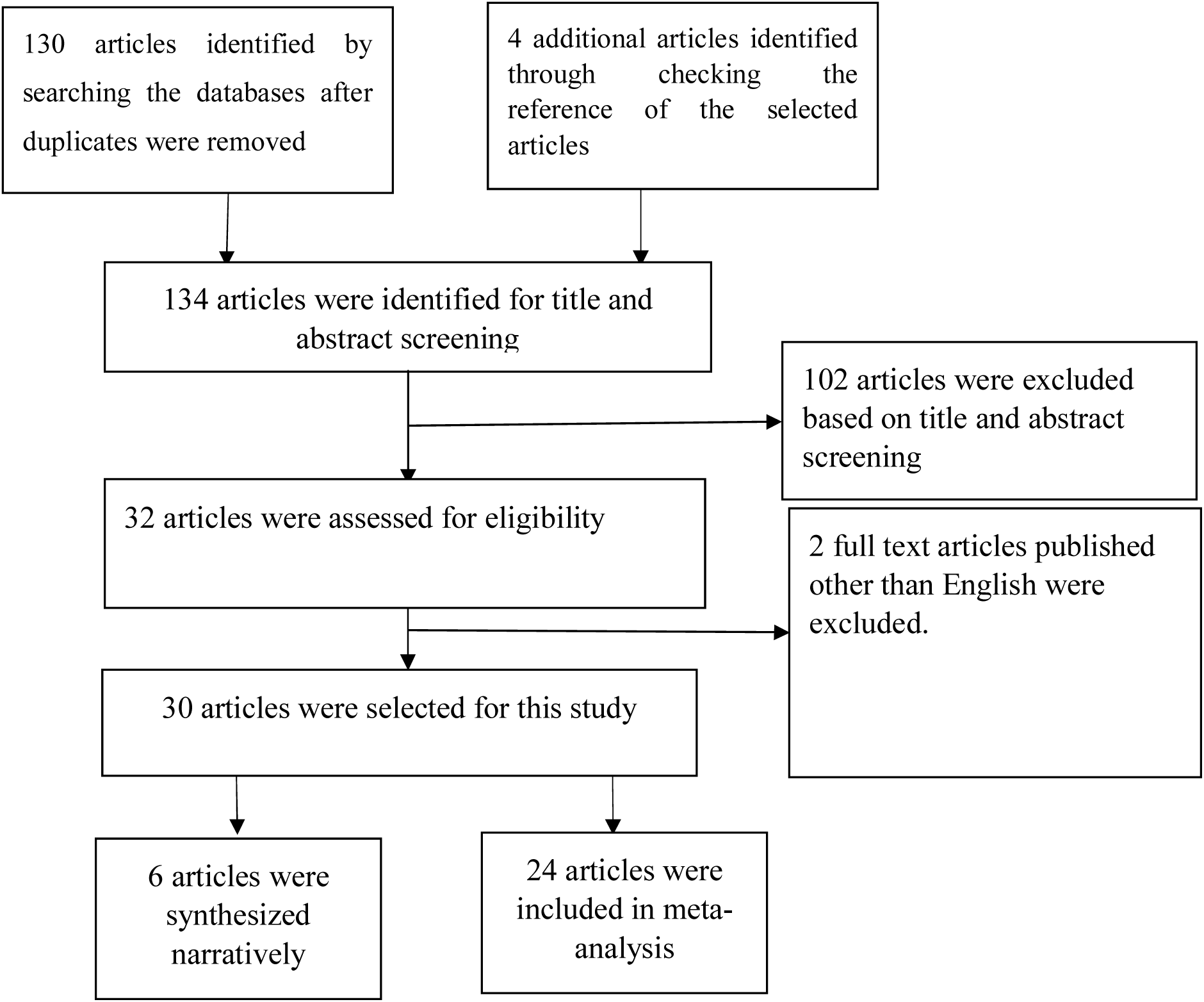
Schematic representation of studies included in the systematic review using the PRISMA checklist and flow diagram

Majority of the studies selected were conducted in China (8) [18,26–32] and its province (6) [33–38]. The remaining studies were conducted in Japan (4) [39–42] followed by South Korea (3) [43–45], Italy (2) [46,47], and France, Germany, and Spain [47]. Four studies included were conducted based on multiple countries’ data [19,48–50].

### Estimated reproduction number

Of the 30 studies included in this review reported different reproductive numbers (Table 1). The estimated reproductive number in this analysis was 2.70 (95% CI, 2.21-3.30) with a very high-heterogeneity (99.5%) (Figure 3). However, we did not find any evidence of publication bias (Supplementary Figure 1). Sub-group analysis was used to address heterogeneity (Table 2, Supplementary Figures 2-5). We found study’s characteristics, such as countries for which the reproductive number estimated, models and methods used to estimate the reproductive number, and the number of cases used to estimate reproductive number were played a significant role of arising such heterogeneity (Table 2). For instance, the estimated reproductive number was around double (R, 4.56, 95% CI, 2.28-9.12) in outside of China than China (R, 2.72, 95% CI, 2.08-3.57). However, in the country level, the highest reproductive number was reported for France (R, 6.32, 95% CI, 5.72-6.98) following Germany (R, 6.07, 95% CI, 5.51-6.69) and Spain (R, 5.08, 95% CI, 4.50-5.73). South Korea was only country reported <1 reproductive number of COVID-19 (R, 0.76, 95% CI, 0.34-1.70). The higher reproductive number was reported if estimated by the MCMC method (R, 4.18, 95% CI, 1.75-9.93) and by the Epidemic curve model (R, 2.86, 95% CI, 2.39-3.42). The average reproductive number found higher if it was estimated for >3162 cases (R, 2.97, 95% CI, 2.09-4.23) than ≤3162 cases (R, 2.50, 95% CI, 1.91-3.28).

**Table 1:**
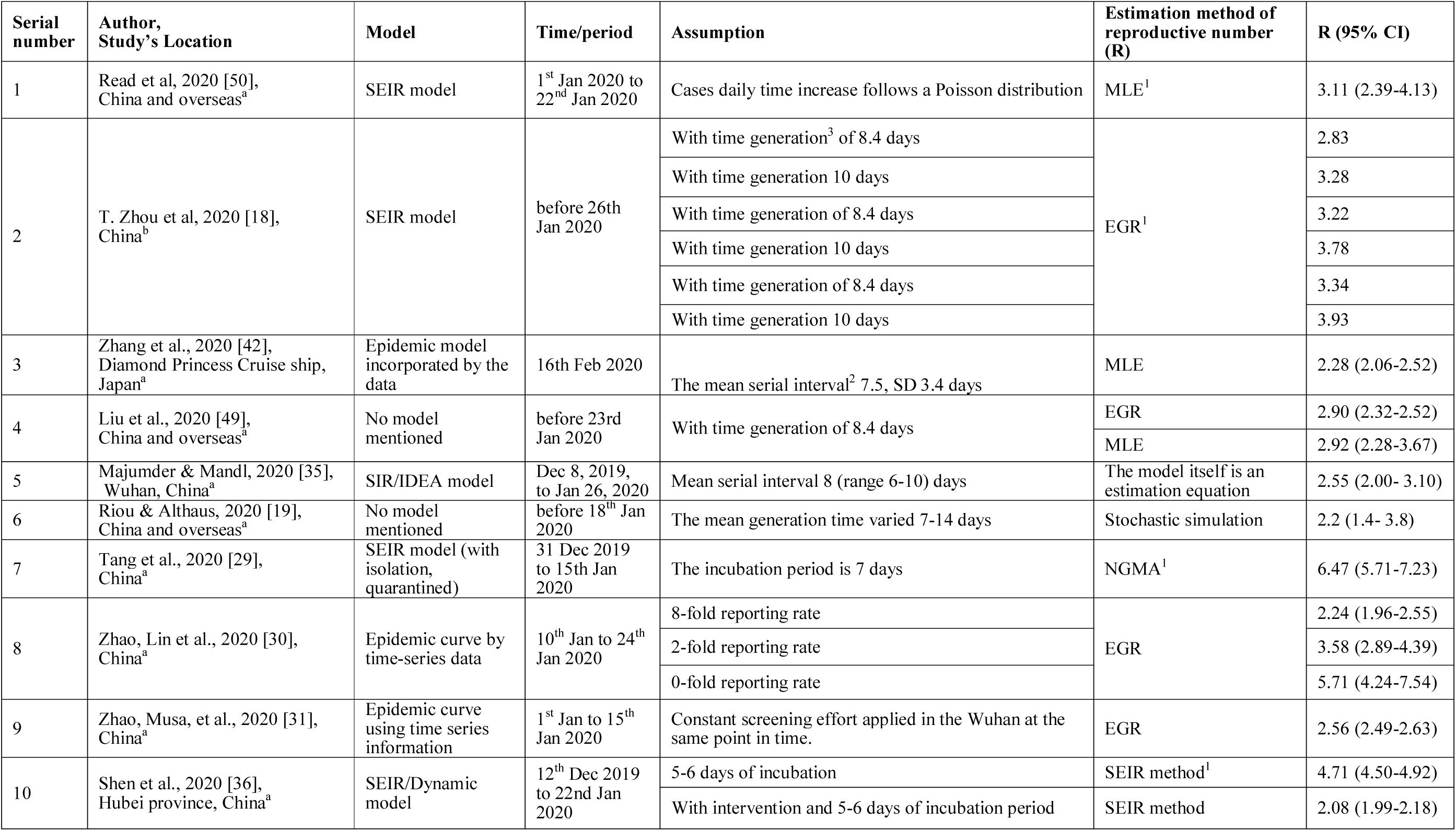
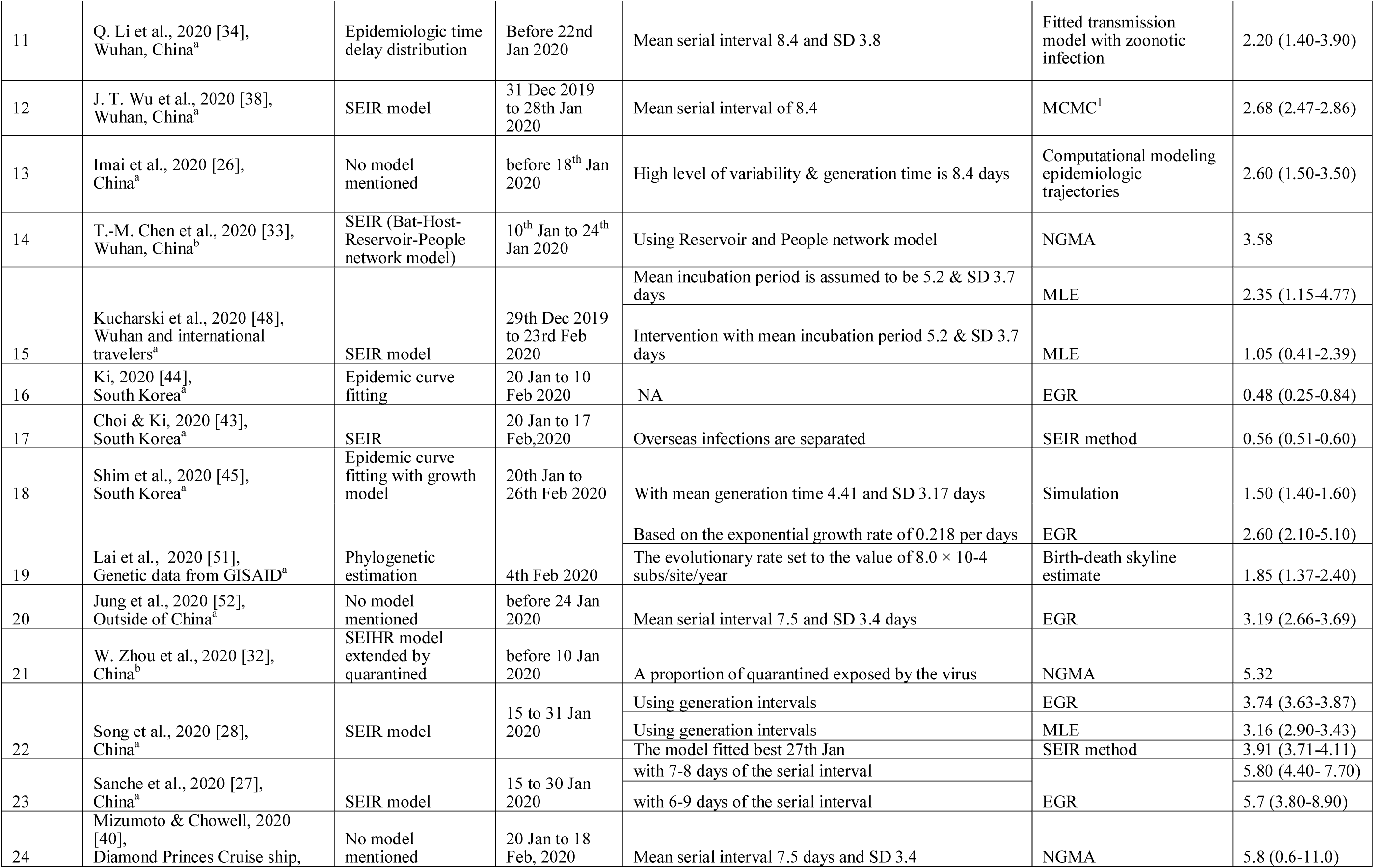
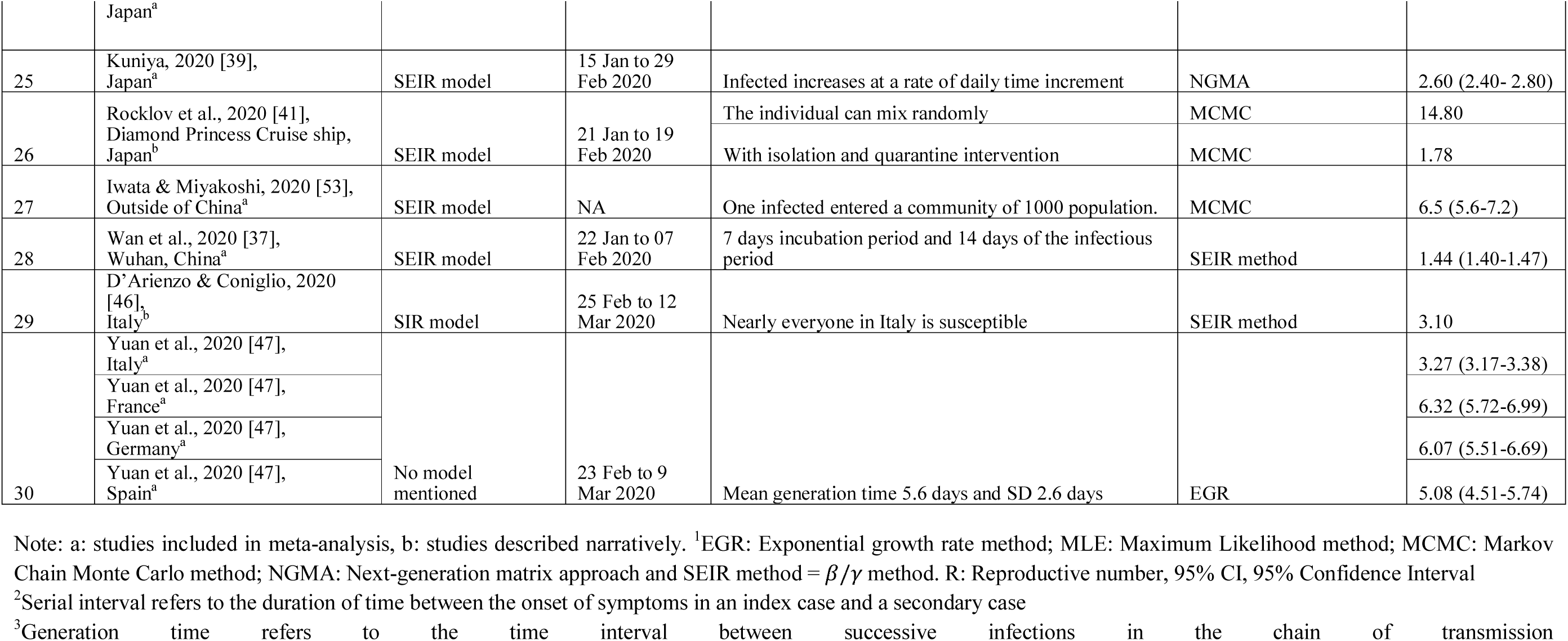
Background characteristics of the studies included in this review.

**Figure 3:**
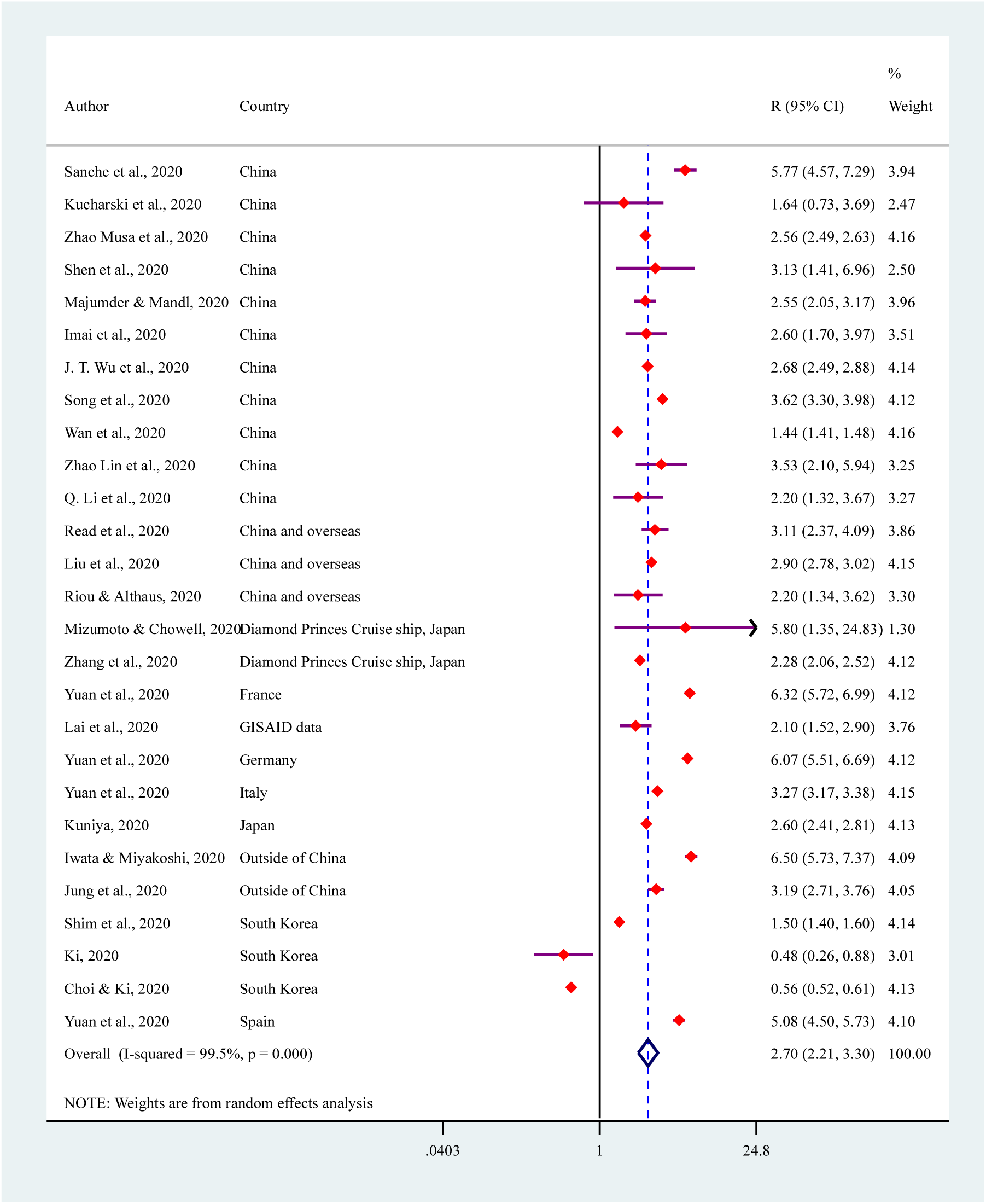
Summarized reproductive number of COVID-19 (total 24 studies with 27 times report of COVID-19’s reproductive number [one study (Yuan et al., 2020) reported estimates for four different countries])

**Table 2:** Sources of heterogeneity of the estimated COVID-19’s reproductive number.

| Characteristics | Number of studies** | R (95% CI) | P |  |
| --- | --- | --- | --- | --- |
|  |  |  | Heterogeneity | Meta-regression |
| **Country** |  |  |  |  |
| China | 11 | 2.72 (2.08-3.57) | <0.01 | <0.01 |
| China and overseas | 3 | 2.90 (2.78-3.02) | 0.490 |  |
| Outside of China | 2 | 4.56 (2.28-9.12) | <0.01 |  |
| Japan | 1 | 2.60 (2.41-2.81) | NA |  |
| Diamond Princes Cruise ship, Japan | 2 | 2.71 (1.33-5.26) | 0.290 |  |
| South Korea | 3 | 0.76 (0.34-1.70) | <0.01 |  |
| Italy | 1 | 3.27 (3.16-3.38) | NA |  |
| Germany | 1 | 6.07 (5.51-6.69) | NA |  |
| Spain | 1 | 5.08 (4.50-5.73) | NA |  |
| France | 1 | 6.32 (5.72-6.98) | NA |  |
| Global Initiative on Sharing AI Influenza Data | 1 | 2.1 (1.52-2.90) | NA |  |
| **Method considered** |  |  |  |  |
| MLE | 4 | 2.63 (2.17-3.18) | <0.01 | <0.05 |
| EGR | 6 | 3.67 (2.90-4.64) | <0.01 |  |
| SEIR | 4 | 1.67 (0.82-3.38) | <0.01 |  |
| MCMC | 2 | 4.18 (1.75-9.93) | <0.01 |  |
| NGMA | 2 | 2.76 (1.82-4.16) | 0.280 |  |
| Others | 6 | 2.11 (1.60-2.79) | <0.01 |  |
| **Model considered** |  |  |  |  |
| SEIR model | 10 | 2.57 (1.69-3.92) | <0.01 | 0.9470 |
| SIR model | 1 | 2.55 (2.05-3.18) | NA |  |
| Epidemic curve | 13 | 2.86 (2.39-3.42) | <0.01 |  |
| **Number of cases** |  |  |  |  |
| ≤3162 | 16 | 2.50 (1.91-3.28) | <0.01 | 0.7881 |
| >3162 | 11 | 2.97 (2.09-4.23) | <0.01 |  |

Note: \*\* Number of studies 24 with reproductive number record 27 time (one study reported estimate for four different countries)

The results of the narrative synthesis are presented in Table 3. Total of six studies were narratively synthesized. The findings of these six studies also supported our summary estimate. A study conducted for Diamond Princes Cruise Ship, Japan found reproductive number of COVID-19 was 14.8 for the period of 21 January to 19 February 2020 [41]. However, this estimated reproductive number was conditioned for not to applied any preventive intervention and infected person can mix randomly to the non-infected persons. When preventive interventions applied this number was reduced to 1.78.

**Table 3:** Narrative synthesis of the studies included in in review.

| Author, Study's Location | Model | Time/period | Assumptions and method | Results |
| --- | --- | --- | --- | --- |
| T. Zhou et al, 2020 [18], China<sup>b</sup> | SEIR model | before 26th Jan 2020 | With time generation of 8.4 and 10 days and using the exponential growth rate method | Estimated basic reproductive number was varied from 2.83 to 3.34 (for 8.4 days generation time) or 3.28 to 3.93 (for 10 days generation time). |
| Tang et al., 2020 [29], China<sup>b</sup> | SEIR model (with isolation, quarantined) | 31 Dec 2019 to 15th Jan 2020 | The incubation period was 7 days, ignoring the asymptomatic infection in the model and using the next generation matrix approach | The estimated reproductive number was 6.47 (5.71-7.23) during the control measures of isolation and quarantine are implementing. |
| T.-M. Chen et al., 2020 [33], Wuhan, China<sup>b</sup> | SEIR (Bat-Host-Reservoir-People network model) | 10<sup>th</sup> Jan to 24<sup>th</sup> Jan 2020 | Assuming the mean incubation 5.2, mean infectious period 5.8 and using the next generation matrix approach | The basic reproduction number estimated was 2.30 from reservoir to person. It was increased to 3.58 when reached person-to-person level transmission. |
| W. Zhou et al., 2020 [32], China<sup>b</sup> | SEIHR model extended by quarantined | before 10 Jan 2020 | Parameterizing cumulative cases, deaths, daily number of media reports and proportion of quarantined exposed by the virus and the estimation method was next generation matrix approach | The basic reproductive number was 5.32. |
| Rocklov et al., 2020 [41], Diamond Princess Cruise ship, Japan<sup>b</sup> | SEIR model | 21 Jan to 19 Feb 2020 | The individual can mix randomly, infectious period was 10 days and contact rate was same as early outbreak using the SEIR method. | The basic reproductive number was 14.80 without any intervention by using 79% infected persons in the ship. However, isolation and quarantine before 62.35% infected cases reduce this number to 1.78. |
| D'Arienzo & Coniglio, 2020 [46], Italy<sup>b</sup> | SIR model | 25 Feb to 12 Mar 2020 | Nearly everyone in Italy were considered as susceptible using the general SEIR method | The Ro was 3.10 while the number varies from 2.46 to 3.09 in different region across Italy. |

## Discussion

This review aimed to provide the reproductive number of COVID-19 based on the global level evidence. A total of 30 studies selected for this study of which 24 studies were included in the meta-analysis. Majority of the included studies were conducted in China. The average estimated reproductive number was 2.70 with evidence of higher heterogeneity across the included studies. The sources of heterogeneity were countries for which the reproductive number estimated, models and methods used to estimate the reproductive number, and the number of cases used to estimate the reproductive number.

The average estimated reproductive number was 2.7; which is higher than the WHO’s estimate of 1. 4 to 2.5. However, this estimate is lower than the previous summarized reproductive number of COVID-19 (3.28) [54]. Numerous measures to reduce new infections of COVID-19 such as social distancing, and controlling international travels are associated with such reduction [17,55]. However, our estimated reproductive number is still very high that leads an exponential increase of new infections. Moreover, the estimated number is still very higher than previous rounds of COVID-19’s like infectious diseases, such as SARS and MERS if we considered time period between the when was estimation done and infections was initially detected. For instance, the reproductive number of SARS and MERS were reduced to 0.95 (95% CI, 0.61-1,23) and 0.91 (95% CI, 0.36-1.44) after 3^rd^ generation of the infection [56]. There are numerous reasons for such a higher reproductive number. First, biological facts of the infection rate and duration of contagion are important to explain such higher reproductive number instead of strict control measures that placed to reduce new infections [57]. For instance, a person could be infected in numerous ways, such as gets physically contacted with the infected person or through environmental transmission by respiratory droplets [58]. Moreover, COVID-19 infected patients may not show symptomatic characteristics up to two weeks of infection. This pre-symptomatic stage is another source to increase new infections exponentially as in this period an infected person is usually confounded in the community with other people. This risk is further increased significantly for the country where population density is high [59].

This study also found evidence of the very high (99.5%) heterogeneity of the estimated reproductive number. Along with the factors described above, characteristics used to estimate reproductive numbers are important source for such heterogeneity. For instance, the reproductive number found higher for the countries where no restriction was applied or restriction was applied in delayed. The forms of restrictions were control people’s movement, personal hygiene, and wearing mask [10,60]. These implications act to control virus transmission from an infected to the susceptible and reduce the new infections. These also affect the average transmissibility of COVID-19 within the specific population and settings [61,62].

Estimation models, assumptions applied, and estimation processes were empirical sources of variability of the estimated COVID-19’s reproductive number [63]. For instance, studies included in this analysis were followed assumption of generation time (which is followed by the gamma distribution) or serial interval (which is followed by the poison distribution) which is an important source of heterogeneity [64–66]. The reason of such difference is the underlying concept: generation time refers to the average time between transmission the virus from an infected person to the non-infected person whereas serial interval refers duration between onset of symptoms in an index case to the transmission in a secondary case [64,65,67]. Moreover, the estimated reproduction number generated by mathematical models is dependent on numerous decisions made by the researcher such as homogeneity or heterogeneity of the population considered; use a deterministic or stochastic approach and which distributions to be used to describe the probable values of parameters [57].

This study was first of its kind that provides an estimation of reproductive numbers based on worldwide’ literature. Moreover, we have considered the heterogeneity of the reproductive numbers estimated worldwide and explored the sources of heterogeneity across selected studies’ characteristics. However, many other factors may explain the sources of heterogeneity of the reported reproductive number of COVID-19 worldwide. We did not explore these because of the lack of data.

## Conclusion

The average estimated reproductive number was 2.70. We found evidence of higher heterogeneity of the reproductive number reported worldwide. There are numerous causes of such heterogeneity, however, study related characteristics were countries for which the reproductive number estimated, methods and models used to estimate reproductive number, and the number of cases considered to estimate reproductive number. This analysis indicates possibility to significant increase of COVID-19 infections in the coming days. Strengthing existing preventive measures as well as new policies and programs are important to reduce new infections.

## Data Availability

Data available on request to the corresponding author

## Acknowledgment

The authors are grateful to the authors of the paper included in this review.

## Funds

This study did not received any financial support.

## Ethical approval

Ethical approval was not necessary for this kind of study.

Abbreviations
COVID-19: Novel Coronavirus 2019
EGR: Exponential Growth Rate
MCMC: Markov Chain Monte Carlo
MERS: Middle East Respiratory Syndrome
MLE: Maximum Likelihood Estimation
SARS: Severe Acute Respiratory Syndrome
SEIR: Susceptible, Exposed, Infected and Removed/Recovered
SIR: Susceptible, Infected and Removed/Recovered

---

**Supplemental Table 1.**
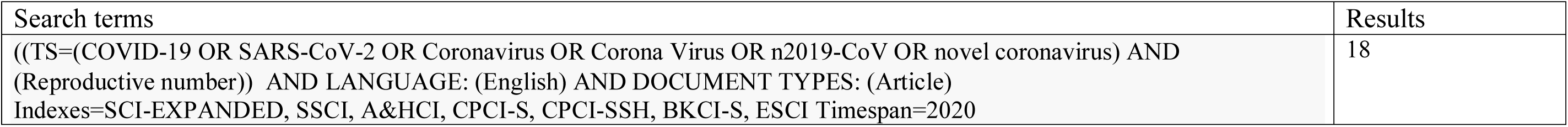
Web of science search results for pre-existing morbidities among COVID-19 patients.

**Supplemental Table 2.**
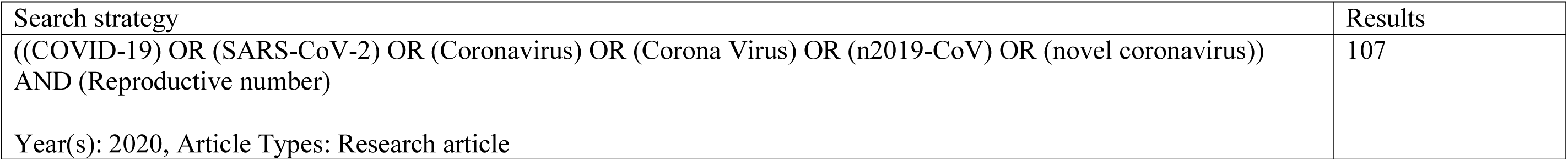
Science Direct search results for reproductive number of COVID-19.

**Supplemental Table 3.**
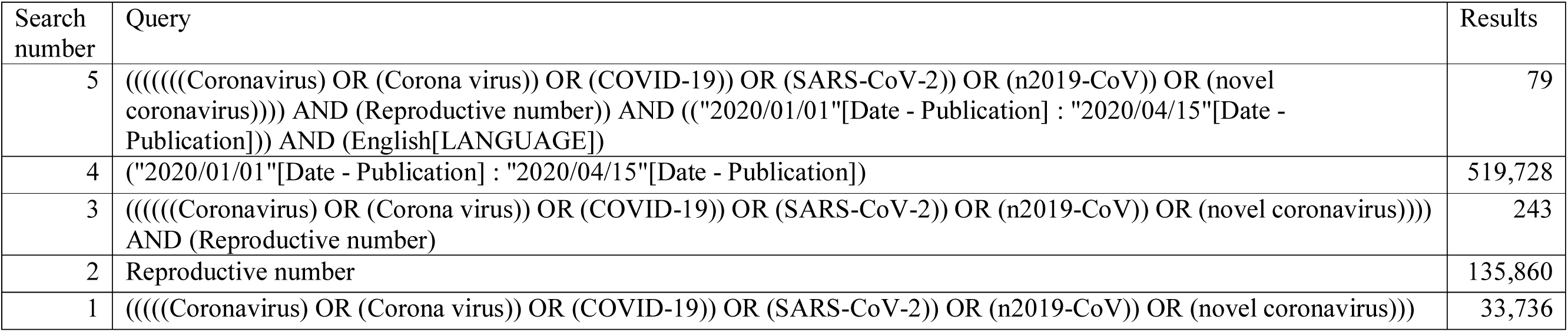
PUBMED search results for reproductive number of COVID-19.

**Supplemental Table 4.** Assessment of the included study through the NIH assessment tool.

| Author | Clarity in objective clearly | Study population clearly and fully described with case definition | The cases consecutive | The subjects comparable | The intervention clearly described | The outcome measures clearly defined, valid, reliable and implemented consistently across all the participants | The length of follow up adequate | The statistical methods well described | The results well described | Total score |
| --- | --- | --- | --- | --- | --- | --- | --- | --- | --- | --- |
| Read, et al | Y | Y | Y | Y | Y | O | N | Y | Y | 7 |
| Zhou et al | Y | Y | Y | Y | Y | Y | N | Y | Y | 8 |
| Zhang et al | Y | Y | Y | Y | O | Y | N | Y | Y | 7 |
| Liu et al | Y | Y | Y | O | Y | Y | Y | Y | Y | 8 |
| Majumder and Mandl | N | Y | Y | Y | N | Y | N | Y | Y | 6 |
| Riou and Althaus | Y | Y | Y | Y | N | Y | Y | Y | Y | 8 |
| Tang et al. | Y | Y | Y | Y | Y | Y | Y | Y | Y | 9 |
| Zhao et al. | Y | Y | Y | Y | O | Y | N | Y | Y | 7 |
| Zhao et al | Y | Y | Y | Y | N | Y | Y | Y | Y | 8 |
| Shen et al | Y | Y | Y | Y | Y | Y | N | Y | Y | 8 |
| Li et al. | Y | Y | Y | Y | N | Y | Y | Y | Y | 8 |
| Wu et al | Y | Y | Y | Y | Y | Y | Y | Y | Y | 9 |
| Imai et al | Y | Y | Y | Y | O | Y | N | Y | Y | 7 |
| Chen et al | Y | Y | Y | Y | Y | Y | N | Y | Y | 8 |
| Kucharski et al. | Y | Y | Y | Y | Y | Y | Y | Y | Y | 9 |
| Ki et al., | Y | Y | Y | Y | Y | Y | Y | Y | Y | 9 |
| Choi and Ki | Y | Y | Y | Y | Y | Y | Y | Y | Y | 9 |
| Shim et al | Y | Y | Y | Y | Y | Y | N | Y | Y | 8 |
| Lai et al., | Y | Y | Y | Y | Y | Y | Y | Y | Y | 9 |
| Jung et al | Y | Y | Y | Y | Y | Y | Y | Y | Y | 8 |
| Zhou et al | Y | Y | Y | Y | Y | Y | Y | Y | Y | 9 |
| Song, et al | Y | Y | Y | Y | O | O | O | Y | Y | 6 |
| Sanche et al. | Y | Y | Y | Y | N | Y | N | Y | Y | 7 |
| Mizumoto and Chowell | Y | Y | Y | Y | Y | Y | Y | Y | Y | 9 |
| Kuniya | Y | Y | O | Y | O | Y | N | Y | Y | 6 |
| Rocklov, et al | Y | Y | Y | Y | Y | Y | Y | Y | Y | 9 |
| Iwata and Miyakoshi | Y | Y | N | Y | N | Y | N | Y | Y | 7 |
| Wan et al | Y | Y | Y | Y | O | Y | Y | Y | Y | 8 |
| D'Arienzo, Coniglio | Y | Y | Y | Y | N | N | N | Y | Y | 6 |
| Yuan, et al. | Y | Y | Y | Y | Y | Y | Y | Y | Y | 9 |
| Yes= Y, No= N, Others (NA, NR, CD)= O |  |  |  |  |  |  |  |  |  |  |

**Supplementary Figure 1:**
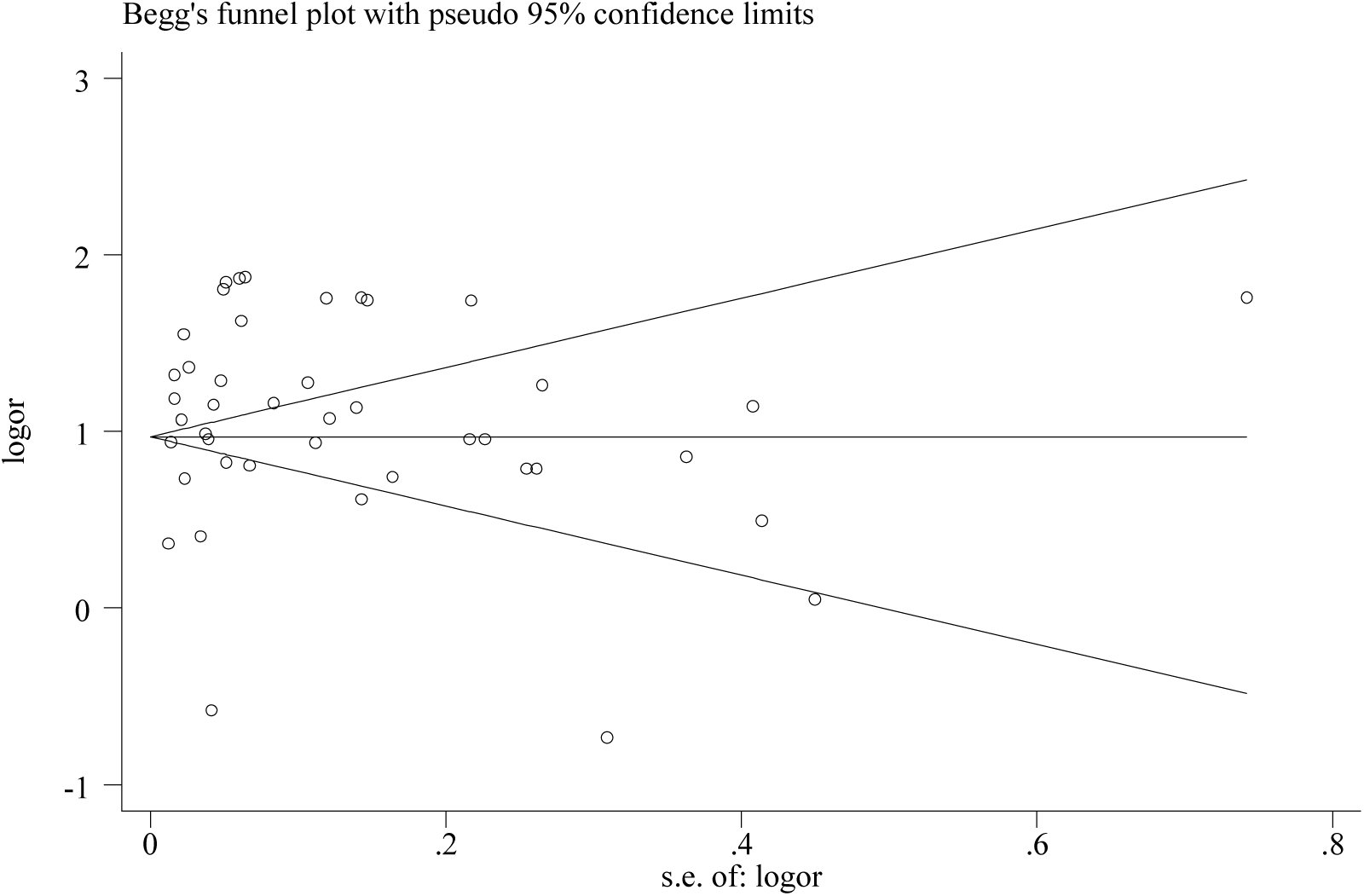
Funnel plot for the included studies estimated reproductive number of COVID-19 to identify publication bias.

**Supplementary Figure 2:**
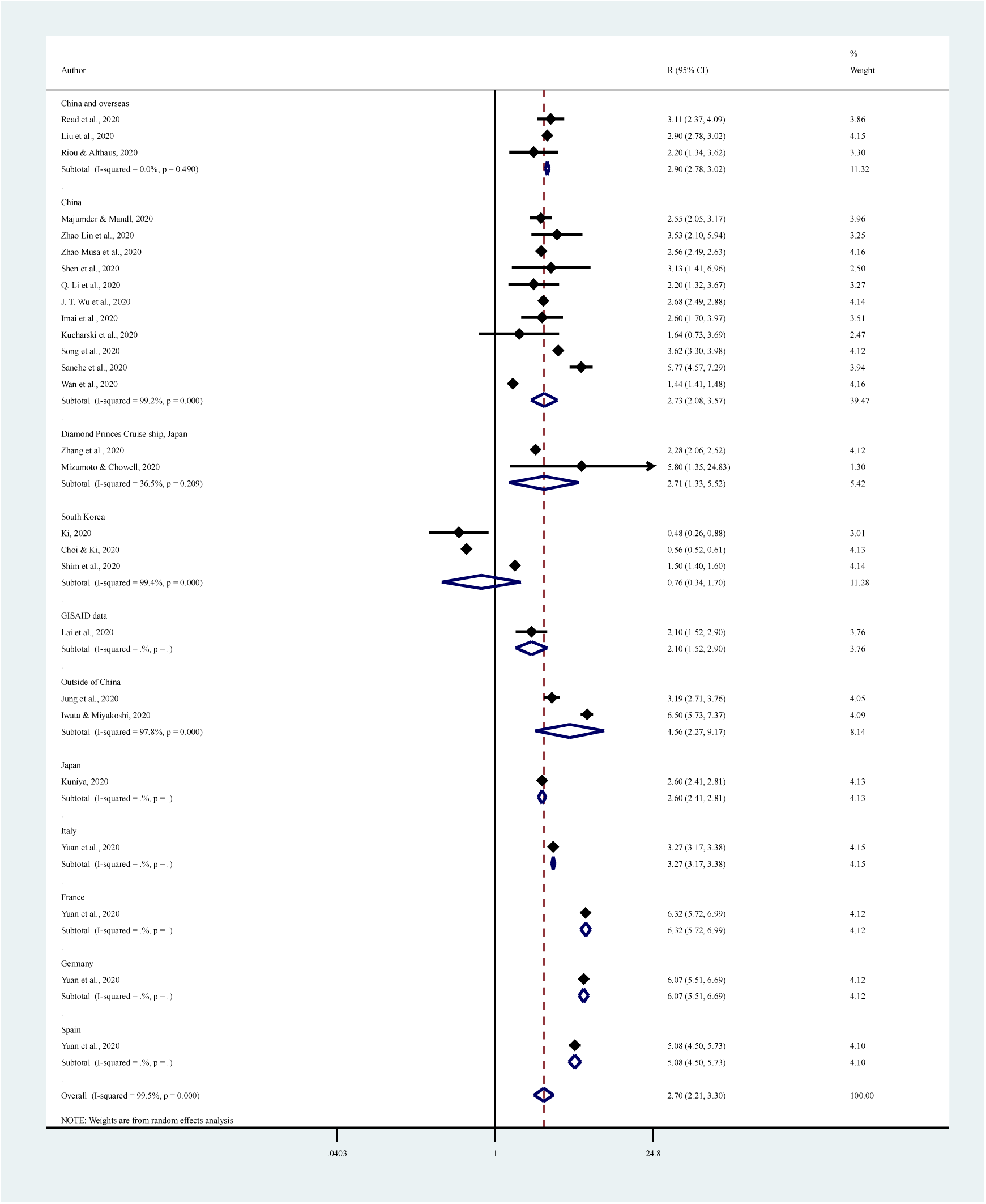
Estimated reproductive number of COVID-19 across countries.

**Supplementary Figure 3:**
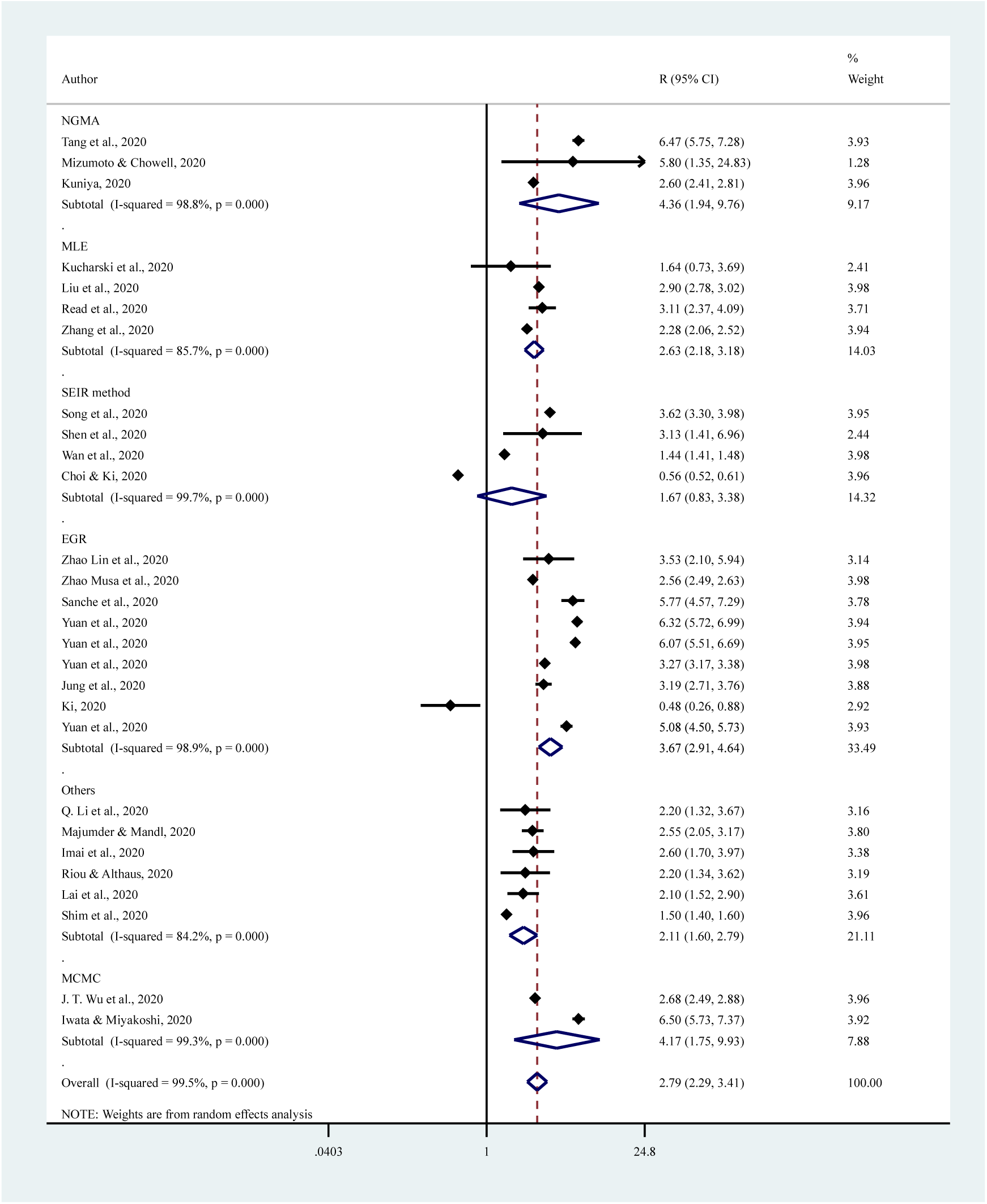
Estimated reproductive number of COVID-19 across methods used to estimate.

**Supplementary Figure 4:**
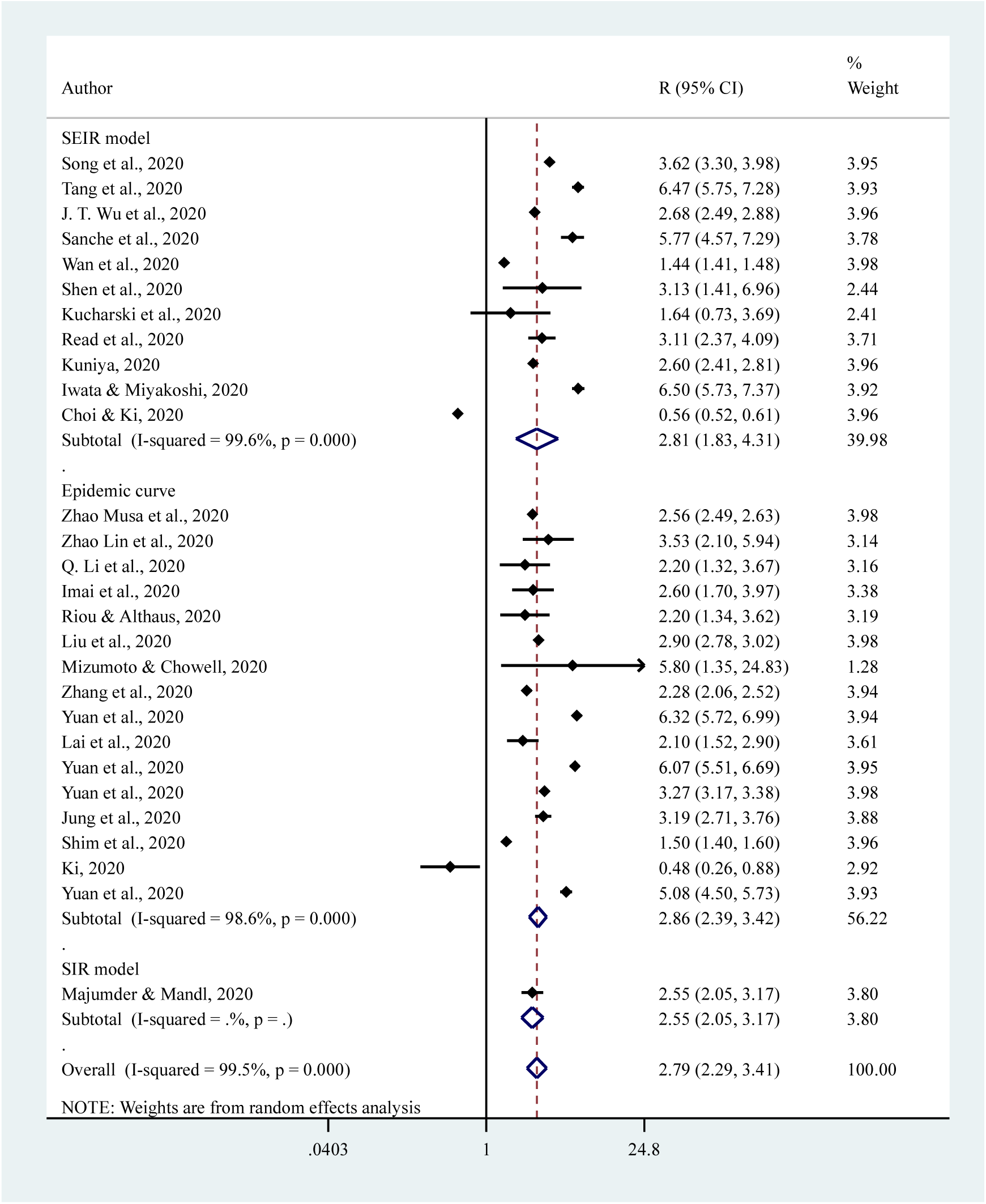
Estimated reproductive number of COVID-19 across models used to estimate.

**Supplementary Figure 5:**
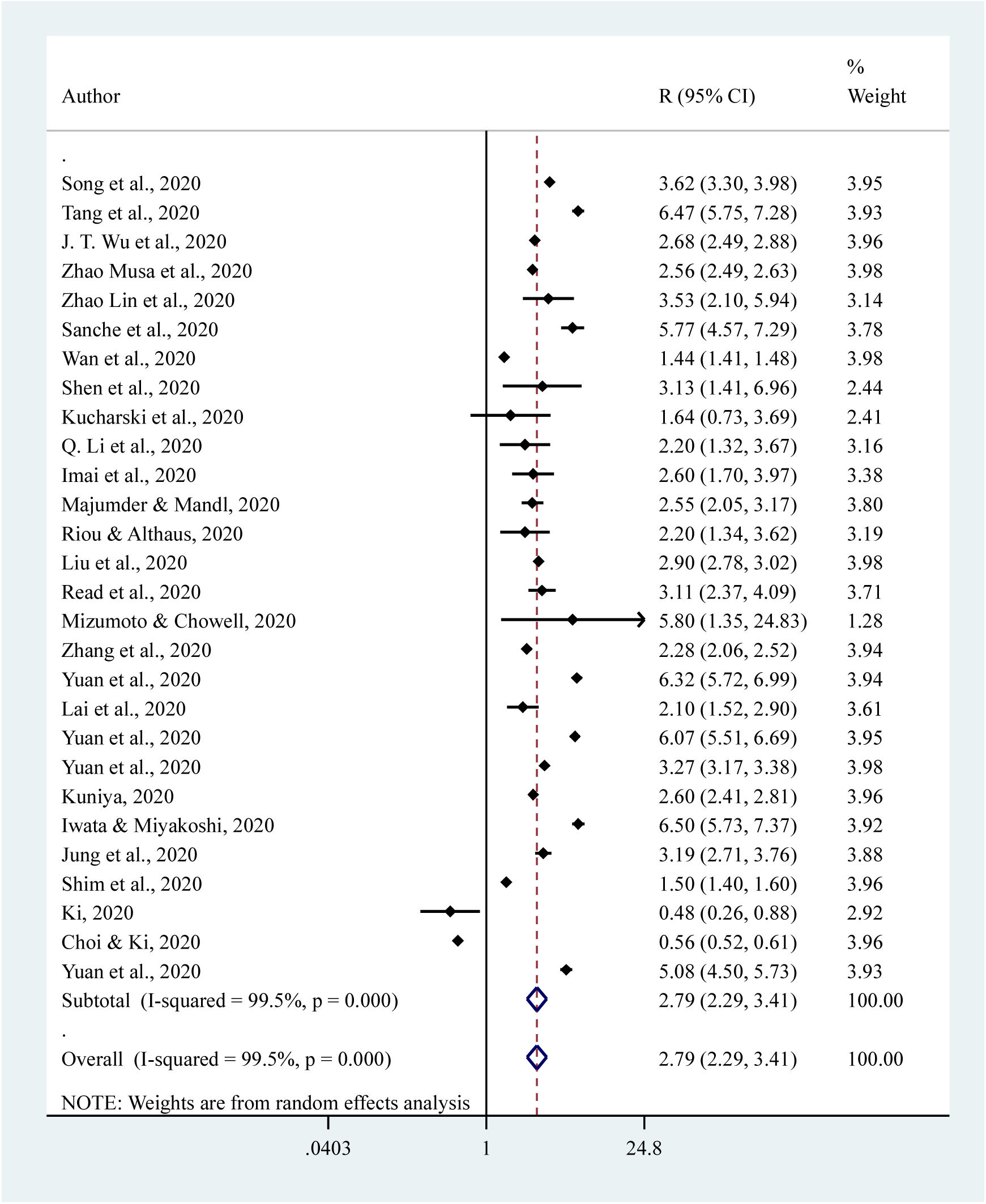
Estimated reproductive number of COVID-19 across the number of cases considered to estimate.

| Section/topic | # | Checklist item | Reported on page # |
| --- | --- | --- | --- |
| **TITLE** |  |  |  |
| Title | 1 | Identify the report as a systematic review, meta-analysis, or both. | 2 |
| **ABSTRACT** |  |  |  |
| Structured summary | 2 | Provide a structured summary including, as applicable: background; objectives; data sources; study eligibility criteria, participants, and interventions; study appraisal and synthesis methods; results; limitations; conclusions and implications of key findings; systematic review registration number. | 2 |
| **INTRODUCTION** |  |  |  |
| Rationale | 3 | Describe the rationale for the review in the context of what is already known. | 4 |
| Objectives | 4 | Provide an explicit statement of questions being addressed with reference to participants, interventions, comparisons, outcomes, and study design (PICOS). | 4 |
| **METHODS** |  |  |  |
| Protocol and registration | 5 | Indicate if a review protocol exists, if and where it can be accessed (e.g., Web address), and, if available, provide registration information including registration number. | 6 |
| Eligibility criteria | 6 | Specify study characteristics (e.g., PICOS, length of follow-up) and report characteristics (e.g., years considered, language, publication status) used as criteria for eligibility, giving rationale. | 7 |
| Information sources | 7 | Describe all information sources (e.g., databases with dates of coverage, contact with study authors to identify additional studies) in the search and date last searched. | 7 |
| Search | 8 | Present full electronic search strategy for at least one database, including any limits used, such that it could be repeated. | 7 |
| Study selection | 9 | State the process for selecting studies (i.e., screening, eligibility, included in systematic review, and, if applicable, included in the meta-analysis). | 7 |
| Data collection process | 10 | Describe method of data extraction from reports (e.g., piloted forms, independently, in duplicate) and any processes for obtaining and confirming data from investigators. | 8 |
| Data items | 11 | List and define all variables for which data were sought (e.g., PICOS, funding sources) and any assumptions and simplifications made. | 8 |
| Risk of bias in individual studies | 12 | Describe methods used for assessing risk of bias of individual studies (including specification of whether this was done at the study or outcome level), and how this information is to be used in any data synthesis. | 8 |
| Summary measures | 13 | State the principal summary measures (e.g., risk ratio, difference in means). | 8 |
| Synthesis of results | 14 | Describe the methods of handling data and combining results of studies, if done, including measures of consistency (e.g., $I^2$) for each meta-analysis. | 8 |

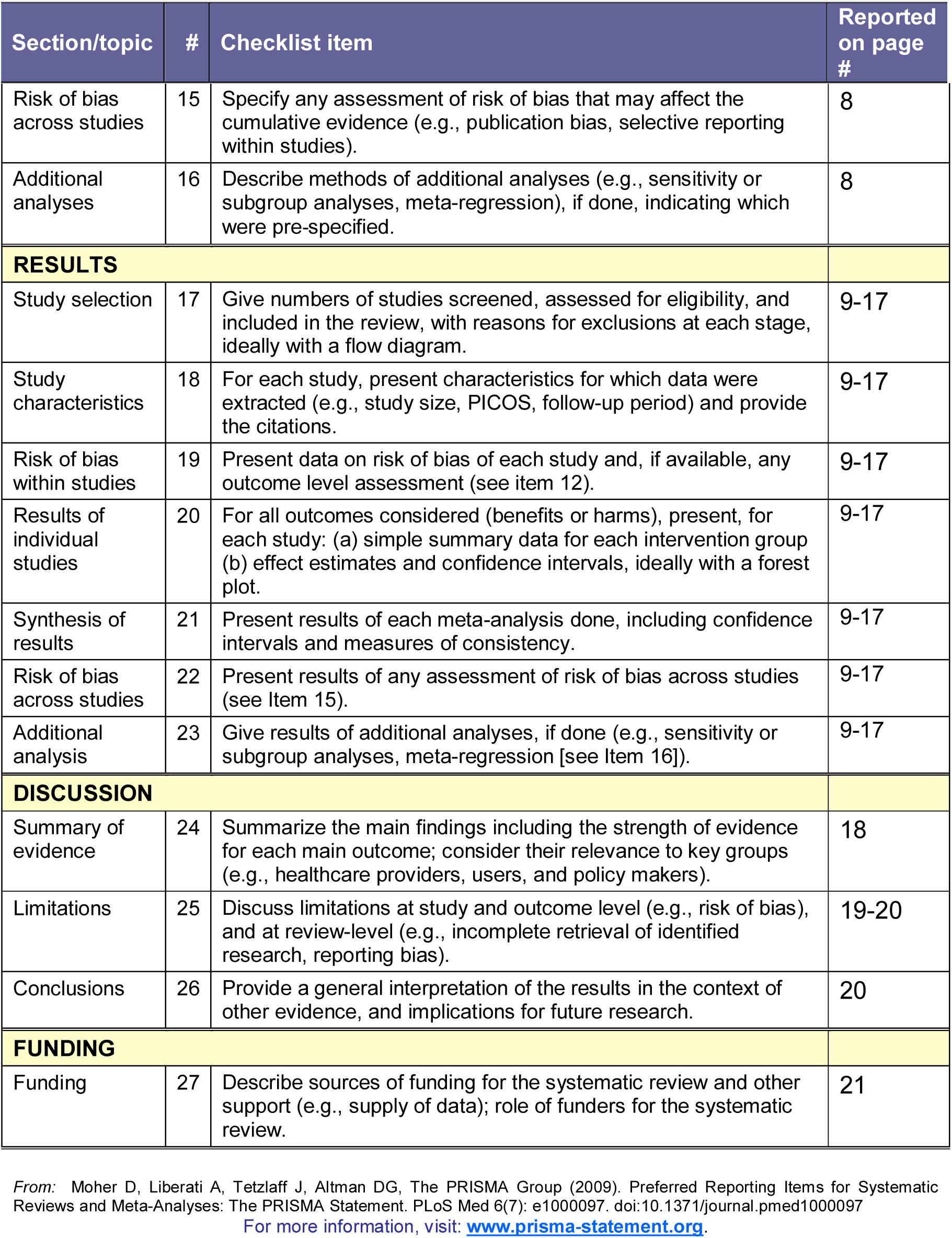

